# Systematic review and meta-analysis of veterinary-related occupational exposures to hazards

**DOI:** 10.1101/2020.09.02.20186775

**Authors:** OO Adebowale, OG Fasanmi, B.B Awosile, MO Afolabi, FO Fasina

## Abstract

**Objective:** Understanding hazards within the veterinary profession is critical for developing strategies to ensure health and safety in the work environment. This study was conducted to systematically review and synthesize data on reported risks within veterinary workplaces.

**Methods:** A systematic review of published data reporting occupational hazards and associated risk factors were searched within three database platforms namely PubMed, Ebscohost, and Google scholar. To determine the proportion estimates of hazards and pooled odds ratio, two random-effects meta-analysis were performed.

**Results:** Data showed veterinarians and students were at high risk of exposure to diverse physical, chemical, and biological hazards. For the biological, chemical and physical hazards, the pooled proportion estimates were 17% (95% CI: 15.0–19.0, p < 0.001), 7.0% (95% CI: 6.0–9.0%, p < 0.001) and 65.0% (95% CI: 39.0–91.0%, p < 0.001) respectively. A pooled odds ratio indicated the odds of physical (OR = 1.012, 95% CI: 1.008–1.017, p < 0.001) and biological exposures (OR = 2.07, 95% CI: 1.70–2.52, p < 0.001) increased more when working with or in contact with animals than non-contact.

**Conclusions:** This review has provided a better understanding of occupational health and safety status of veterinarians and gaps within the developing countries. Veterinarians including students are at considerable risk of occupational-related hazards. The need to improve government and organisation policies and measures on occupational health and safety is therefore crucial, most importantly in Africa.

## 1. Introduction

Occupational hazards are work-related injuries or illnesses people are exposed to at the workplaces, and these hazards contribute immensely to the global burden of diseases [1]. Globally, approximately 2.3 – 2.7 million workers die from work-related injuries or illnesses annually [2,3], and total loss due to this worldwide, has been estimated to be about $2.99 trillion annually [1,3].

The veterinary profession is comprised of a diverse group of individuals who interact with a wide variability of animal species under a working environment that creates exposure to injuries [4]. In the United States (US), the profession has been ranked as the fifth-highest industry for the incidence of non-fatal work related injuries. This is not far behind police and fire protection services, while public health hospitals were not in the top 20 [5]. Although, some reports showed exposure to work-related hazards are more common in the developed than developing countries [6], whether this is a true assessment is doubtful and indeterminate because most African countries lack occupational health and safety standards, implementations, risks assessment and adequate reporting protocols.

The reported processes of injuries in the profession included physical hazards such as needlestick injuries (NSI), animal bites, kicks, scratches and crushing by equipment used for animal restraint, which imposed physical harm or hurt to individuals [5,7]. Other occupational threats include exposure to biological (especially zoonoses), and chemical hazards (high doses of radiation and pesticides) which increase the risk of birth defects in offspring of female veterinarians [8]. Scientific data have also disclosed seroprevalence against different zoonoses is greater among veterinarians than in the general population, suggesting that veterinarians could act as sentinels to detect emergent diseases and propagators of infections [6]. Common zoonoses found primarily associated with health risks or illnesses among veterinary students and professionals were caused primarily by bacteria, parasites, viruses and fungi [6].

Therefore, this work aims to assess available pieces of evidence regarding exposures to zoonoses, physical and chemical threats among professionals and students in the veterinary discipline, and synthesize the associated risk factors.

## 2. Methodology

### 2.1 Study Type and Search approach

A systematic literature review and meta-analysis was conducted to identify scientific articles documenting hazards, investigate risk factors or practices and association with exposures. Preferred Reporting Items for Systematic Reviews and Meta-Analyses Statement (PRISMA) was used to summarize the article selection process. To select important papers systematic search within PubMed, Ebscohost and Google scholar database was conducted. The search strategy included the key terms “occupational hazards”, “zoonoses”, “veterinary students”, “veterinarians”, “risk factors”, which were combined with the Boolean operator “AND” “OR”. Paper selections were based on information provided in the titles and/or abstracts.

### 2.2 Eligibility criteria for the selection of relevant materials

Only papers published in English were eligible, and no restrictions were placed on the location of studies, except time (2007 – 2017). The following inclusion criteria for selection were used: 1). Paper title and abstract addressed the questions of interest i.e. “occupational health hazards report on veterinary professionals and students in the past 10 years (2007 – 2017) and document the associated risk factors or practices 2). Papers should be cross-sectional observational studies, cohort, case-control studies relevant to research focus 3). Studies providing association data either univariable or multivariable analyses were included. 4). Confirmed zoonoses reports using laboratory detection methods. Mining and filtering of articles were carried out based on set inclusion criteria, and the selected papers were scrutinized critically to find the best available pieces of evidence. Paper abstracts that reported intervention studies, reviews, systematic reviews and meta-analysis, and conference proceedings were excluded.

Two of the authors independently read and examined all titles and abstracts of papers identified in the search, retrieved and selected articles that focused on and met the inclusion criteria. During full-text study final selection, duplicated studies were removed and approval from both authors was obtained for a paper to be included.

### 2.3 Data extraction and analysis

Data extracted onto Excel spreadsheet (Microsoft Office Package 2016; Microsoft Corporation, Redmond, USA) included: 1) year of publication, 2) Authors, 3) location of study, 4) study design, 5) study population, 6) identified hazards, 7) laboratory methods, 8) prevalence and, 9) associated risk factors. Initial descriptive statistics were carried out to summarize the various data retrieved. Then random-effects meta-analysis was carried out to allow for any heterogeneity between studies. Two random-effects meta-analysis was done, firstly, to calculate the pooled (weighted) proportions with respective 95% confidence intervals for the different types of occupational hazards (i.e. biological, physical, and chemical hazards) among veterinarians. Second meta-analysis was done to calculate the pooled (weighted) ratio measures (Odds ratio or relative risk) of risk factors associated with occupational hazards. The risk factor meta-analysis was only done for working or contact with animals as a risk factor for occupational hazards among veterinarians. This is premised on sufficient published articles reporting ratio measures for working or contact with animals compared with other reported risk factors. The pooled prevalence and associated study estimate, and pooled Odds ratio and study estimates were presented using forest plots. The *I^2^* statistic (a measure of inconsistency) was used to assess the variation between studies due to heterogeneity. A value of 0% shows no observed heterogeneity; increasing values indicate increasing heterogeneity. The *I^2^* statistic with cutoff values ≤25%, 26-≤50%, and ≥75% was considered as low, moderate, and substantial heterogeneity. Subgroup analysis was performed to account for potential sources of heterogeneity between studies. Statistical significance was set at P< 0.05 while statistical analysis was carried out using STATA SE/15.0 (College Station, Texas 77845 USA).

## 3. RESULTS

### 3.1 Literature search outcome

The preliminary search retrieved 5,258 articles and a total of 33 articles were retained for the review following paper filtering for eligibility. Rejections were mainly based on the unavailability of either the risk association or prevalence data or microbiological confirmation methods. These 33 articles cut across various territories such as Africa, Switzerland, Europe, Asia, South and North America, the United States, and the United Kingdom (figure 1). Summary of the article selection process presented in figure 2.

**Figure 1:**
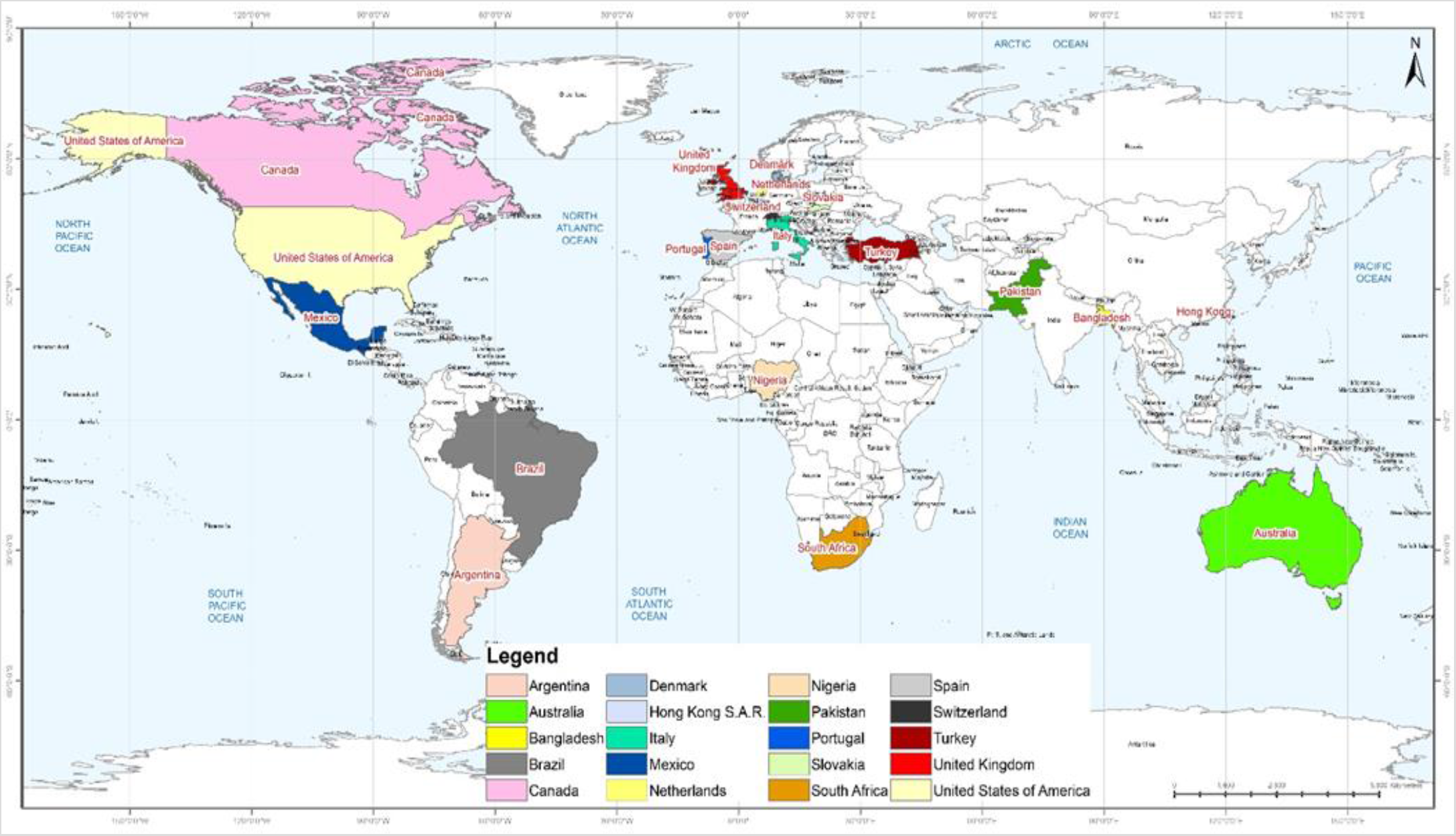
Spatial distribution of countries from which information on occupational hazards and exposure among the veterinary profession were retrieved.

**Figure 2.**
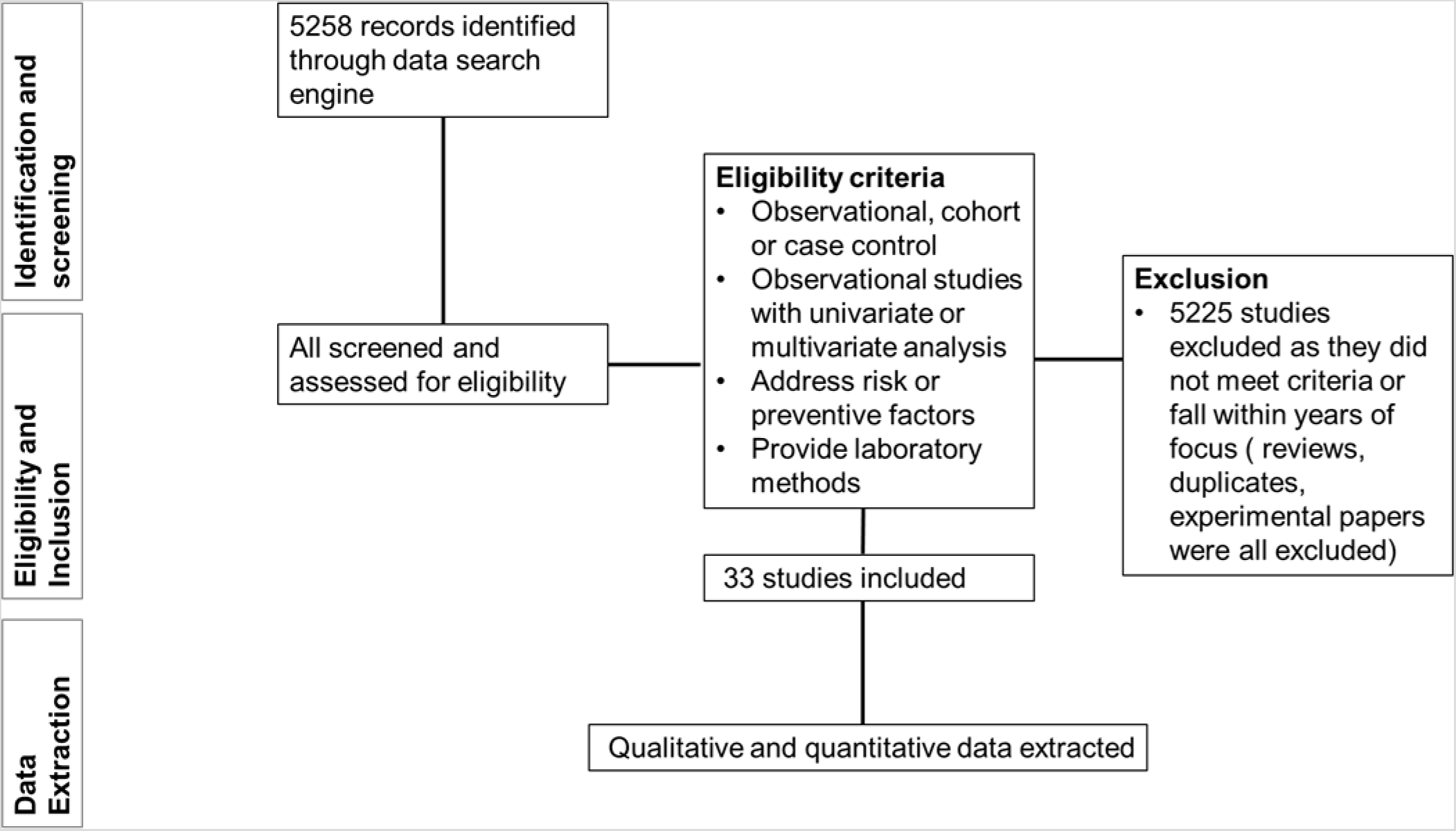
Flow chart for literature selection.

### 3.2 Characteristics of included published articles

Included studies originated from Africa (n = 2), Asia (n = 4), Australia/Oceania (n = 4), Europe (n = 12), North America (n = 8), and South America (n = 3). Of the 33 papers included, twenty – six(78.8%, 95% CI [ 61.9 – 89.62]) were cross-sectional, five (15.1%, 95% CI [6.2 – 31.4]) case-control, and two (6.1%, 95% CI [0.7 – 20.6]) cohort studies. The target populations for the various studies include: 1) Veterinarians (57.6%, 95% CI [40.8 – 72.3]), 2) Veterinary students (9.1%, 95% CI [2.4 - 24.3]), 3) Both Veterinarians and Veterinary students (9.1%, 95% CI [2.4 – 24.3]), and 4) Veterinarians, veterinary students and others (24.4%, 95% CI [12.6 – 41.3]). Twenty – five articles(75.8%, 95% CI [ 58.8 – 87.4]) described exposures to varieties of biological hazards mainly zoonoses, two chemical (6.1%, 95% CI [0.7 – 20.6]), three physical (9.1%, 95% CI [2.4 – 24.3]) and three (9.1%, 95% CI [2.4 – 24.3]) described all exposures. A total of 10 works (30.3%) employed serology only, 5 (15.1%) culture and molecular, 3(9.1%) culture and serology, 2 articles each (6.1%) molecular methods only, and culture, serology and molecular detection techniques respectively. Table 1 further showed the characteristics of papers included in the review.

**Table 1.**
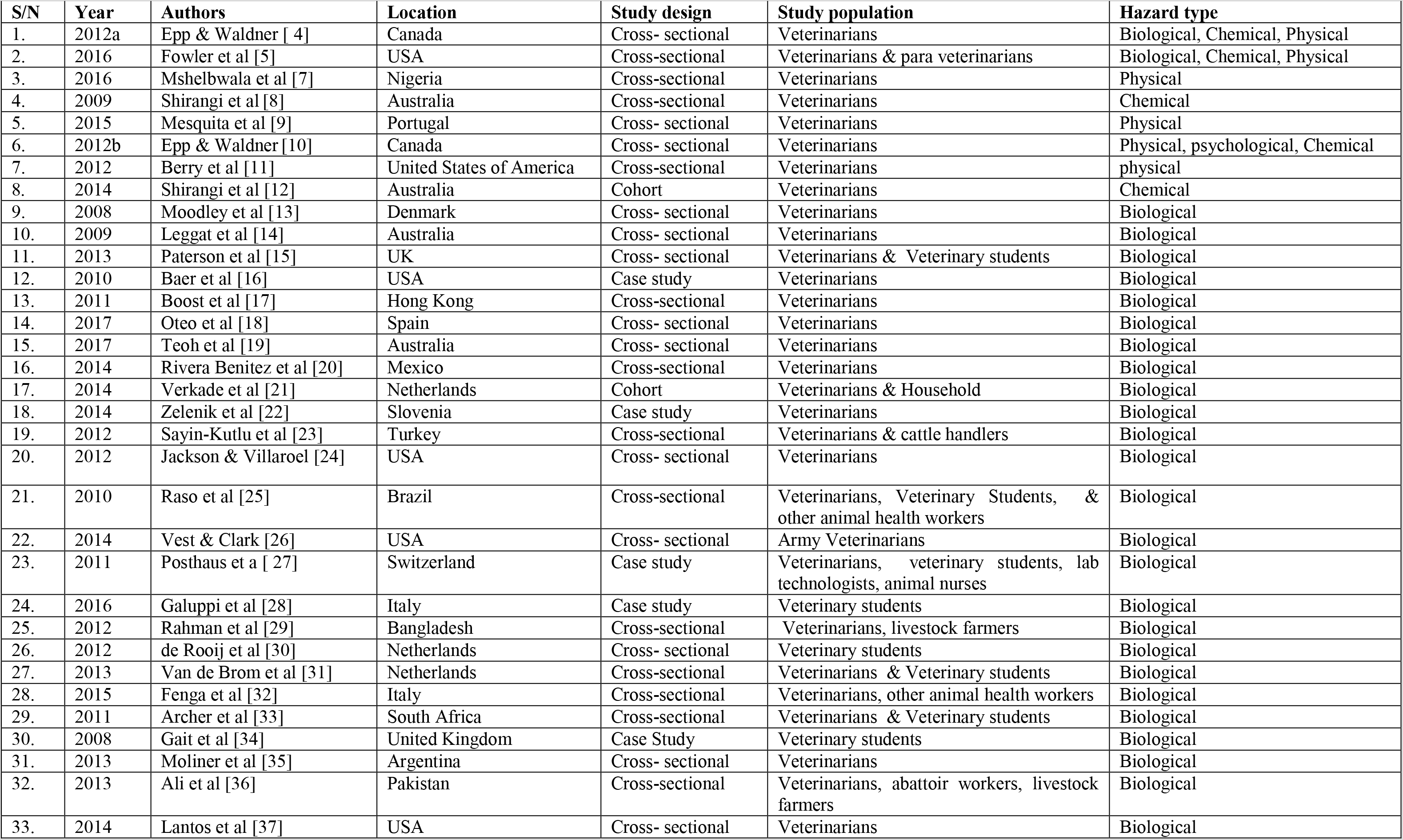
Summary of identified article characteristics according to year, authors, location, study design, study population, and hazard types.

### 3.3 Hazard exposure and risk factors

#### 3.3.1 Physical and psychological hazards

The most common physical hazard identified was needlestick injury (NSI), followed by traumatic injuries and stress. High prevalence exposure to needlestick injuries (range: 65.0 – 79.5%) among the veterinary practice was commonly reported [4, 5,9,7]. Being a male subject (OR 2.8, 95% CI 1.4– 6.0, and working with poultry daily (OR 2.4, 95% CI 1.1–6.2) were found to be significantly associated with NSI in a study conducted in Nigeria [7]. Another study reported veterinarians working in small animal practice especially with dogs were more likely to have experienced an NSI [9]. Furthermore, veterinary technicians were significantly (P < 0.001) more likely than veterinarians to report NSI since their training include recapping needles at school or workplace, a study in the USA reported [5]. Besides NSI, exposure to animal bites was found to be common among companion animal and mixed animal veterinarians than the equine or food animal practitioners [4].

Furthermore, moderate stress and chronic traumatic disorders/injuries were documented among Veterinarians. Western Canadian veterinarians experienced severe (5.2%) to moderate stress (52.6%) related to the workplace [10]. Stress levels were found to be similar for veterinarians in the academia, industry and government but significantly higher (P < 0.001) in female Veterinarians and those who worked more than 40-hour per week (P = 0.001) [10]. Several injuries reported included back pain, limb strains, fall, vehicle accidents, concussion, head injuries and animal bites/scratch. Cumulative traumatic disorders (CTD) were also reported in females more than in male veterinarians, which required treatment or restriction from regular work activities [11]. Large animal practitioners were found more likely to report a CTD than other veterinary practitioners, and the injuries were more often around the shoulders, forearms, elbows, hands, or knees [11] with resultant one or more days off work [10]. No physical or psychological risk exposures were reported for veterinary students in the articles reviewed. Table 2 describes all physical and psychological hazards and associated risk factors.

**Table 2.**
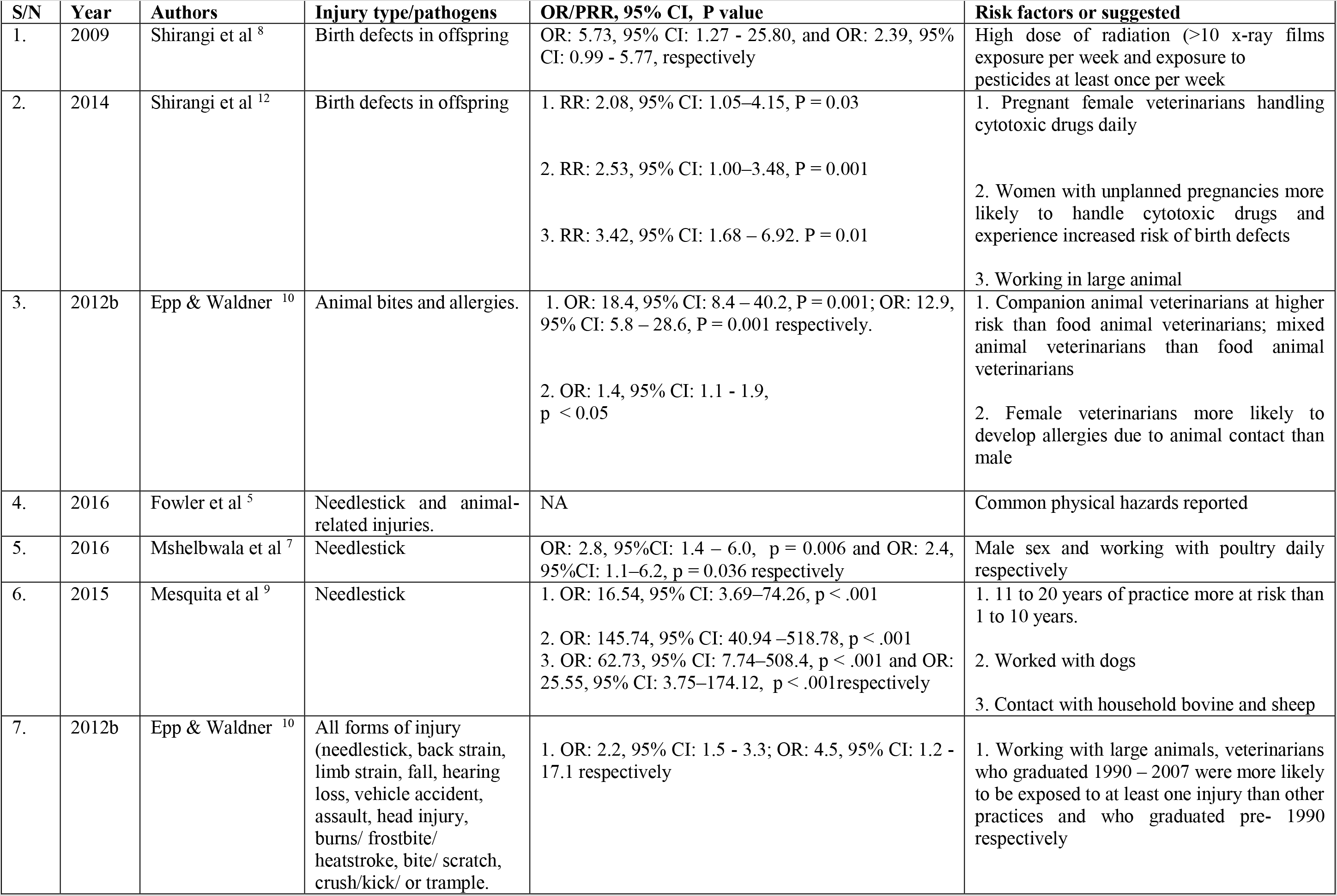
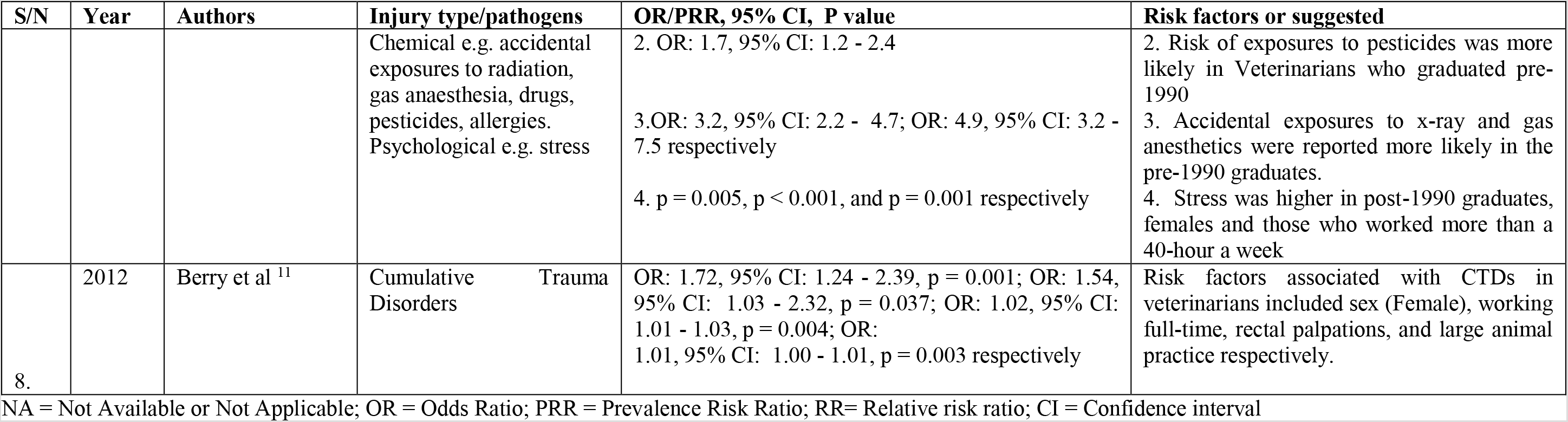
A description of all physical, chemical and psychological hazards identified in the systematic reviews and the associated risk factors/practices.

#### 3.3.2 Chemical hazards

Limited identified chemical hazards and associated risk factors are described in Table 2. Exposure to chemicals occurs in all vet work environment but highest in the private practice [8,10,12]. Veterinarians in this category are more likely to be victims of accidental exposure to drugs (hormones, antimicrobials), sterilizing and cleansing agents, gas or injection anaesthesia and pesticides [10]. A gender-exclusive increased risk of birth defects in offsprings occurred more in female veterinarians working exclusively in small animal practice than in the other environments following exposure to high dose(s) of pesticides at least once per week or in other cases, cytotoxic drugs [8, 12].

#### 3.3.3 Specific biological hazards or Zoonoses common in the veterinary profession

Zoonoses naturally transmitted infections between vertebrate animals and man were recognized as major occupational risks for the veterinary profession due to the degree of animal contact. The most commonly identified zoonoses were methicillin-resistant *Staphylococcus aureus* (MRSA), Q- fever, bartonellosis, cryptosporidiosis, and brucellosis.

##### Methicillin-resistant Staphylococcus aureus (MRSA)

Based on the reviewed literature, MRSA was identified by various diagnostic methods ranging from the traditional culture to molecular typing techniques [13, 15, 17, 21]. The proportions of MRSA colonization in veterinarians varied from 2.6 – 14.7% and veterinarians were six times more at risk of MRSA exposure than non-veterinarians [13]. The direct contact with small animals, cattle or horses predisposed personnel to a higher chance of being MRSA carriers [13]. No exposure was reported among veterinary students

##### Coxiella burnetti

This pathogen of occupational importance causes Q-fever in humans. All the four studies included performed serological assays most especially immunofluorescent assays (IFAT) and Enzyme-linked immunosorbent assay (ELISA). The prevalence of seropositivity to pathogen ranged from 1.4 – 73.7% [4, 24, 26, 30–32]. One of the studies recorded 18.7 % among Dutch veterinary students [30]. Risk factors associated with exposure based on multivariable logistic regression model were identified to include contact with farm animals, students being in advanced year of study, having had exposure to a zoonosis before the study and having ever lived on a ruminant farm [30]. Alternatively, among the Dutch veterinary professionals, the associated risk of exposure was linked with the hours of contact with animal per week, the number of years of post-graduation, being in the rural or suburban residence, being a practising veterinarian, and occupational contact with swine [31] (Table 3).

**Table 3:** A summary of biological hazards and the associated risk factors/practices identified for the systematic review[4][5][13][14][15][16][17][18][19][20][21][22][23][24][25][26][27][28][29][30][31][32][33][34][35][36][37]

| S/N | Year | Authors | Injury type/pathogens | Methods for pathogen confirmation | OR/PR, 95% CI, P value | Risk factors or suggested |
| --- | --- | --- | --- | --- | --- | --- |
| 1. | 2012a | Epp & Waldner [4] | Zoonoses | NA | OR: 8.6, 95% CI: 1.1 - 65.1, P = <0.001 | Mixed animal veterinarians had higher odds of developing a personal zoonosis than others |
| 2. | 2016 | Fowler et al [5] | Infectious diseases (dermatophytosis, bite wound infection, salmonellosis, cryptosporidiosis) | NA | NA | Respondents reported acquiring at least 1 zoonotic infection at some point during their career. |
| 3. | 2008 | Moodley et al [13] | MRSA | Culture and PCR | P=0.02, P<0.001, P<0.001 respectively | MRSA exposure 6 times higher in Veterinarians than non-veterinarian professionals, exposure to small animals, horses respectively |
| 4. | 2009 | Leggat et al [14] | Hand dermatitis (HD) | NA | 1. OR: 4.3, 95% CI: 2.7–6.6, P < 0.001<br>2. OR: 3.5, 95% CI: 2.2–5.4, P < 0.001<br>3. OR: 15.5, 95% CI: 5.4–44.5, P < 0.001 | 1. Veterinarians with a current allergic disease<br>2. In female veterinarians<br>3. Reporting allergies within last 1 year |
| 5. | 2013 | Paterson et al [15] | Methicillin-resistant *Staphylococcus aureus* (MRSA) | Culture, multiplex polymerase chain reaction (PCR), matrix assisted laser desorption/ionization (MALDI-TOF) | NA | Contact with livestock |
| 6. | 2008 | Baer et al [16] | *Leptosira* | Microscopic agglutination test | NA | Contact with an apparently healthy pet rat |
| 7. | 2008 | Boost et al [17] | MRSA | Culture, PCR, multilocus sequence typing (MLST) | P < 0.001, P = 0.03 respectively | Contact with large animals |
| 8. | 2017 | Oteo et al [18] | *Bartonella* | Culture, PCR, DNA sequencing, | NA | Veterinarians working with companion animal |
| 9. | 2017 | Teoh et al [19] | *Rickettsia felis* and *R. typhi* | Indirect micro immunofluorescence antibody testing (MIFAT) | OR: 0.756, 95% CI: 0.582–0.982, P = 0.04; OR: 0.752, 95% CI: 0.579–0.975, P = 0.034 respectively<br>2. OR: 0.611, 95% CI: 0.38–0.982, P = 0.044<br>3. OR: 1.381, 95% CI: 0.973–1.96, P = 0.075 | Older veterinarians > 60 years at reduced risk<br>*R. felis* or generalized *R. felis* or *R. typhi*<br>2. Veterinarians recommending flea treatments<br>3. Veterinarians located at southeastern Australian states of Victoria or Tasmania |
| 10. | 2012 | Rivera Benitez et al [20] | Rubula virus, Encephalomyocarditis virus & *Leptospira* | Hemagglutination inhibition test, viral neutralization test | OR: 1.38, P < 0.05 | Number of visits to farm increased exposure to Encephalomyocarditis virus |
| 11. | 2014 | Verkade et al [21] | MRSA | Culture, (spa) typing and multiple-locus variable-number tandem repeat analysis (MLVA) | PRR: 9.3; 95% CI 2.8 – 38.5; PRR: 2.1; 95% CI 1.0 – 4.6 respectively | Veterinarians with persistent MRSA significantly increased carriage in household members |
| 12. | 2012 | Zelenik et al [22] | *Listeria* | Culture, serology, and pulsed-field gel electrophoresis (PFGE) | NA | Assisted delivery of a stillborn calf |
| 13. | 2010 | Sayin-Kutlu et al [23] | *Bartonella* | Indirect IFA | 1. OR: 5.166; 95% CI, 1.532–17.425, P = 0.008. | 1. Age <35 years' old |
| 14. | 2010 | Jackson & Villaroel [24] | Zoonoses - Rabies, Ringworm, Sarcoptic mange, Campylobacteriosis, Cryptosporidiosis, Giardiasis | NA | 1. P = 0.034; P = 0.031 respectively | 1. Veterinarians below 30 years and recent graduates than experienced veterinarians had reduced risk |
| 15. | 2008 | Raso et al [25] | *Chlamydia psittaci* | Microimmunofluorescence (MIF) | NA | Authors suggested an under-reporting of this disease in Brazil and the need for risk studies |
| 16. | 2012 | Vest & Clark [26] | *Coxiella burnettii* | IFAT | 1. PR: 1.96; 95% CI 1.15–3.35, P = 0.015<br>2. PR: 2.16, 95% CI 0.91–5.1, P = 0.09<br>3. PR: 2.89, 95% CI 1.13–7.4, P = 0.032, and PR: 3.17, 95% CI 1.03–9.71, P = 0.039 respectively | 1. Women veterinarians than the men category<br>2. Veterinarians within 40–49 age group<br>3. Women deployed to Operation Iraqi Freedom |
| 17. | 2011 | Posthaus et al [27] | *Mycobacterium tuberculosis* | IFN-g release assay | NA | This case study reported the potential infection in veterinary personnel with M. tuberculosis due to contact infected dog |
| 18. | 2016 | Galuppi et al [28] | *Cryptosporidium parvum* | Ziehl–Neelsen staining, nested PCR, PCR-restriction fragment length polymorphism analysis (PCR-RFLP) | NA | The case study linked the transmission of the pathogen from foals hospitalized in an equine perinatology unit (EPU) to an outbreak in veterinary students. |
| 19. | 2012 | Rahman et al [29] | *Brucella* | Rose Bengal Test (RBT), Standard Tube Agglutination Test (STAT), iELISA, Q-PCR | 1. OR: 59.8, 95% CI: 6.40 - 559.93, p < 0.001<br>2. OR: 9.9, 95% CI: 1.03 - 95.30, p = 0.047; OR: 14.2, 95% CI: 1.56 - 129.6, p = 0.019 respectively | 1. More likely in handling goats than cattle.<br>2. Duration of contact with animals for 16 – 25 and > 26 years than ≤ 5 years respectively |
| 20. | 2012 | de Rooij et al [30] | *Coxiella burnettii* | Immunofluorescence assay (IFA) | OR: 3.27, 95% CI: 2.14 – 5.02; OR year 6: 2.31, 95% CI: 1.22 – 4.39; OR year 3–5: 1.83, 95% CI : 1.07 – 3.10; OR: 1.74, 95% CI :1.07–2.82; OR: 2.73, 95% CI :1.59 – 4.67 respectively | The study direction; contact with farm animals; advanced year of study (years 6 and 3-5); having had a zoonosis during the study, and ever lived on a ruminant farm respectively |
| 21. | 2013 | Van de Brum et al [31] | *Coxiella burnettii* | Indirect immunofluorescent assay and Enzyme-linked immunosorbent assay (ELISA) | 1. OR: 3.9, 95% CI: 1.5 – 10.1, p = 0.005; OR: 13.1, 95% CI: 4.7 – 36.2, p < 0.001; OR: 26.8, 95% CI: 8.1 – 88.2, p < 0.001 respectively.<br>2. OR: 6.3, 95% CI: 2.6 – 15.1, p < 0.001; OR: 7.9, 95% CI: 3.1 – 20, p < | 1. The number of hours with animal contact per week that is 10 -19 or 20 – 29 or > = 30 hours than < 10 hours.<br>2. Number of years as veterinarian graduate i.e. 3 -13 or 14 – 21 or > 22 years than ≤ 2 years |
|  |  |  |  |  | 0.001; OR: 17.4, 95% CI: 6.0 – 50.8, < 0.001 respectively.<br>3. OR: 17.9, 95% CI: 3.6–88.1, p < 0.001; OR: 11.9, 95% CI: 2.1–68.5, p = 0.037 respectively<br>4. OR: 15.8, 95% CI: 2.9–87.2, p < 0.001<br>5. OR: 3.4, 95% CI: 1.1–10.2, p < 0.001<br>6. OR: 6.3, 95% CI: 3.1 – 12.9, p < 0.001<br>7. OR: 7.0, 95% CI: 1.5 – 31.9, p = 0.004<br>8. OR: 4.7, 95% CI: 2.4 – 9.3, p < 0.001 | 3. Living in a rural or semi urban area<br>4. Livestock veterinarian<br>5. Contact with cows<br>6. Contact with ruminants birth products<br>7. Contact with birth products of pets<br>8. Practicing on cow farm with reports of abortion |
| 22. | 2015 | Fenga et al [32] | *Coxiella burnetii* | ELISA | NA | The study demonstrated a high seroprevalence of *C. burnetii* in animal health workers including veterinarians |
| 23. | 2011 | Archer et al [33] | Rift Valley Virus | ELISA, Real time- PCR (RT-PCR) and/or loop-mediated isothermal amplification assay) and/or virus isolation | RR: 16.3, 95% CI: 2.3 - 114.2 | Performing an animal autopsy |
| 24. | 2008 | Gait et al [34] | *Cryptosporidium parvum* | (PCR-RFLP) and DNA sequencing | NA | A case report of outbreak cryptosporidiosis among veterinary students. The outbreak was linked to poor hand hygiene on a farm with enzootic *C parvum* in calves. |
| 25. | 2013 | Moliner et al [35] | *Brucella*, *Toxoplasma*, *Bacillus anthracis*, *Tricophyton*, *Leptospira*, *Mycobacterium*, others | NA | 1. OR: 4.79, 95% CI: 2.29 –10.01, p = 0.0001; OR: 8.40, 95% CI: 4.28 – 16.48, p = 0.0001 respectively<br>2. OR: 2.08, 95% CI: 1.19 – 3.62, p = 0.0099 | Risk factors identified for brucellosis were:<br>1. The number of years of Practice i.e. 11- 20 and > 20years than <10 years<br>2. Veterinarians working in a zone characterized by a high prevalence of brucellosis and specialized agriculture and milk farms zone |
| 26. | 2013 | Ali et al [36] | *Brucella* | RBT, Serum agglutination test, RT- PCR | OR: 1758.5, 95% CI: 48.47- 63799.12, p < 0.001. | Consumption of raw milk increased exposure to brucellosis |
| 27. | 2014 | Lantos et al [37] | *Bartonella* | Culture, PCR, IFA, DNA sequencing | p = 0.006 | A history of travel to Asia was more common among veterinary subjects |

NA = Not Available or Not Applicable; OR = Odds Ratio; PRR = Prevalence Risk Ratio; RR= Relative risk ratio; CI = Confidence interval

##### Cryptosporidium Parvum

Only two case reports of outbreaks among groups of veterinary students were reported from Italy [28] and the UK [34]. The species were identified in both studies by culture-based methods, PCR-restriction fragment length polymorphism, DNA sequence typing and nested PCR. The outbreak among the Italian veterinary students was plausibly linked with the introduction of *Cryptosporidium* in an equine perinatology unit (EPU) due to an asymptomatic foal [28]. Meanwhile, the outbreak among UK students was documented to be caused by a ‘lapse in hygiene’ on a farm with known infected calves [34].

##### Bartonella species

*Bartonella* species are important emerging pathogens in human and veterinary medicine [37]. High(73.0%) seroprevalence of infection with *Bartonella* spp. was found among companion animal veterinary personnel from Spain in one of the studies reviewed [18]. Meanwhile, the other two reports indicated lower prevalence between 22.0 [23] and 28.0% [37]. A few risk determinants of exposure to pathogen included being less than 35 years of age [23], contact with ticks [23] and history of travel to Asia [37]. Culture, PCR, DNA sequencing and Immunofluorescence assay (IFA) were the main diagnostic techniques used in these studies.

##### Brucella species

The prevalence of infection ranged from 0.1 – 29.1% in our review [29,35,36]. The pathogen was identified mainly by serology and PCR based techniques including the Rose Bengal Test (RBT), Standard Tube Agglutination Test (STAT), indirect enzyme-linked immunosorbent assay (iELISA), and Q-PCR). Contact with animals especially goats, number of years of practice or veterinarians working in a zone characterized by a high prevalence of brucellosis were identified as contributing factors for increased odds of exposure to brucellosis (Table 3). The review also showed the more the number of years accumulated as a graduate, the greater the likelihood of illness [35].

#### 3.3.4 Meta-analysis of proportion estimates of occupational hazards and ratio measure for working or contact with animals as a risk factor for occupational hazards among veterinarians

Based on a meta-analysis, the pooled proportion estimate of biological hazards among veterinarian was 17% (95% CI: 15.0–19.0, p< 0.001). The overall between-study heterogeneity was significant and substantial (*I^2^* = 98.98 %, p < 0.001). Subgroup analysis of different biological hazards was presented in Figure 3, relatively high proportions 31.0% (95% CI: 0.0–62%, P = 0.05), 26.0% (95% CI: 16.0–36.0%, p< 0.001) and 24.0% (95% CI: 0.0–49.0%, p< 0.05) were recorded for bartonellosis, Q-fever and viral infections respectively. However, the estimated proportion for other biological hazards including brucellosis, leptospirosis, cryptosporidiosis, and MRSA was between 7.0% and 14.0% and statistically significant at p< 0.05. For the chemical hazard, the pooled proportional estimate was calculated from two studies with 7.0% (95% CI: 6.0–9.0%, P< 0.001) estimated proportion (Figure 4). However, for physical hazard, a pooled estimate of 65.0% (95% CI: 39.0–91.0%, p< 0.001) was calculated. The overall between-study heterogeneity was significant and substantial (*I^2^* = 99.71 %, p < 0.001). The subgroup analysis returned an estimated proportion of 75.0% (95% CI: 68.0–82.0%, p< 0.001) for needle stick injury among veterinarians and 25% (95% CI: 23.0–27.0%, p< 0.001) for cumulative trauma disorders among the veterinarians (Figure 5). Meta-analysis was done for only working or contact with animals as a risk factor for occupational hazards among veterinarians (Figure 6). The pooled odds ratio for working or contact with animals was OR = 1.013 (95% CI: 1.008–1.017, p< 0.001) with overall between-study heterogeneity significant and substantial (*I^2^* = 99.7 %, p < 0.001). Subgroup analysis returned a pooled odds ratio of OR = 1.012 (95% CI: 1.008–1.017, p< 0.001) for working or contact with animals as a risk factor for physical hazards among veterinarian. However, the pooled odds ratio for working or contact with animals as a risk factor for biological hazards among veterinarian was OR = 2.07 (95% CI: 1.70–2.52, p< 0.001).

**Figure 3:**
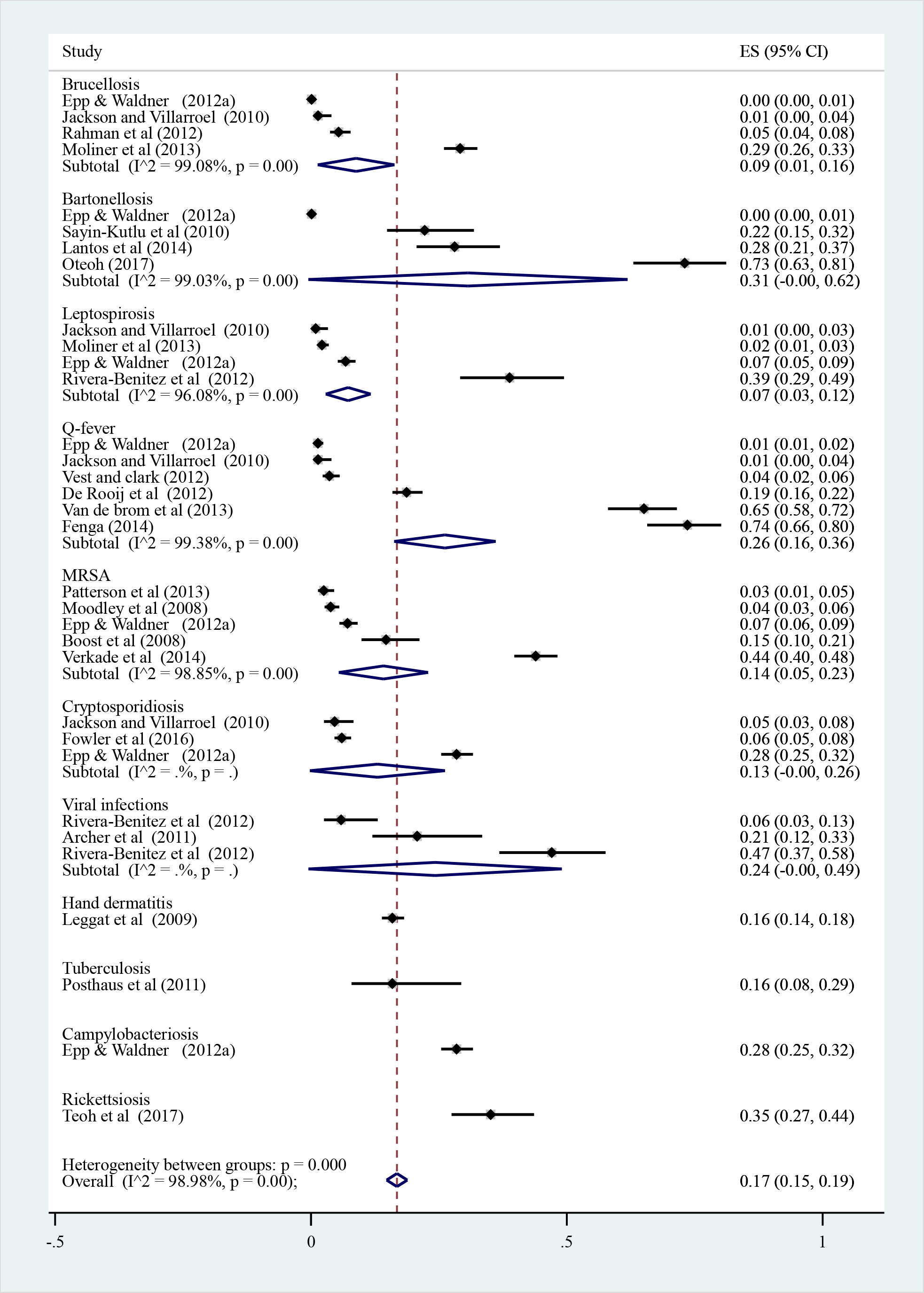
Forest plot of pooled prevalence of biological hazards among veterinarians

**Figure 4:**
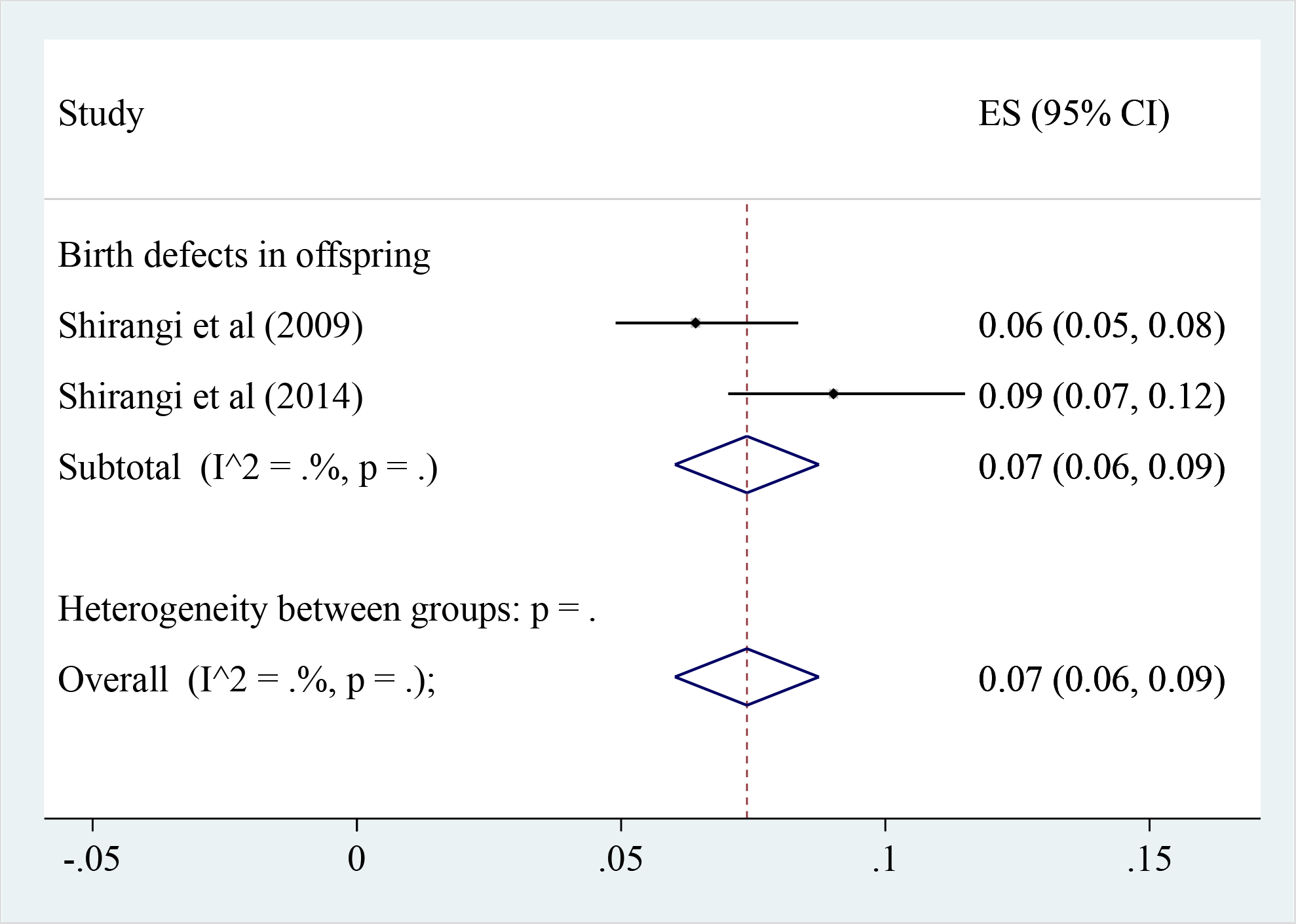
Forest plot of pooled prevalence of chemical hazards among veterinarians

**Figure 5:**
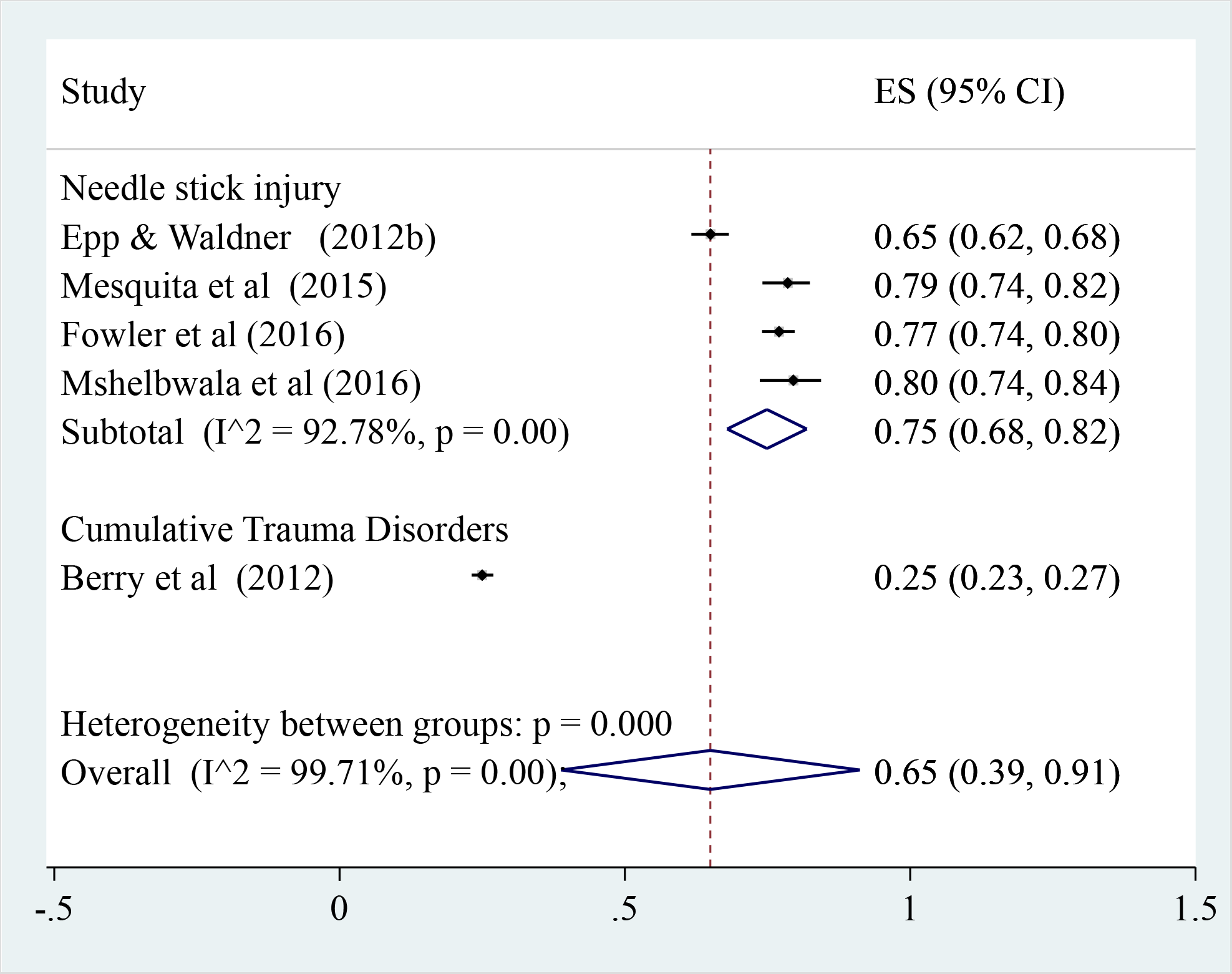
Forest plot of pooled prevalence of physical hazards among veterinarians

**Figure 6.**
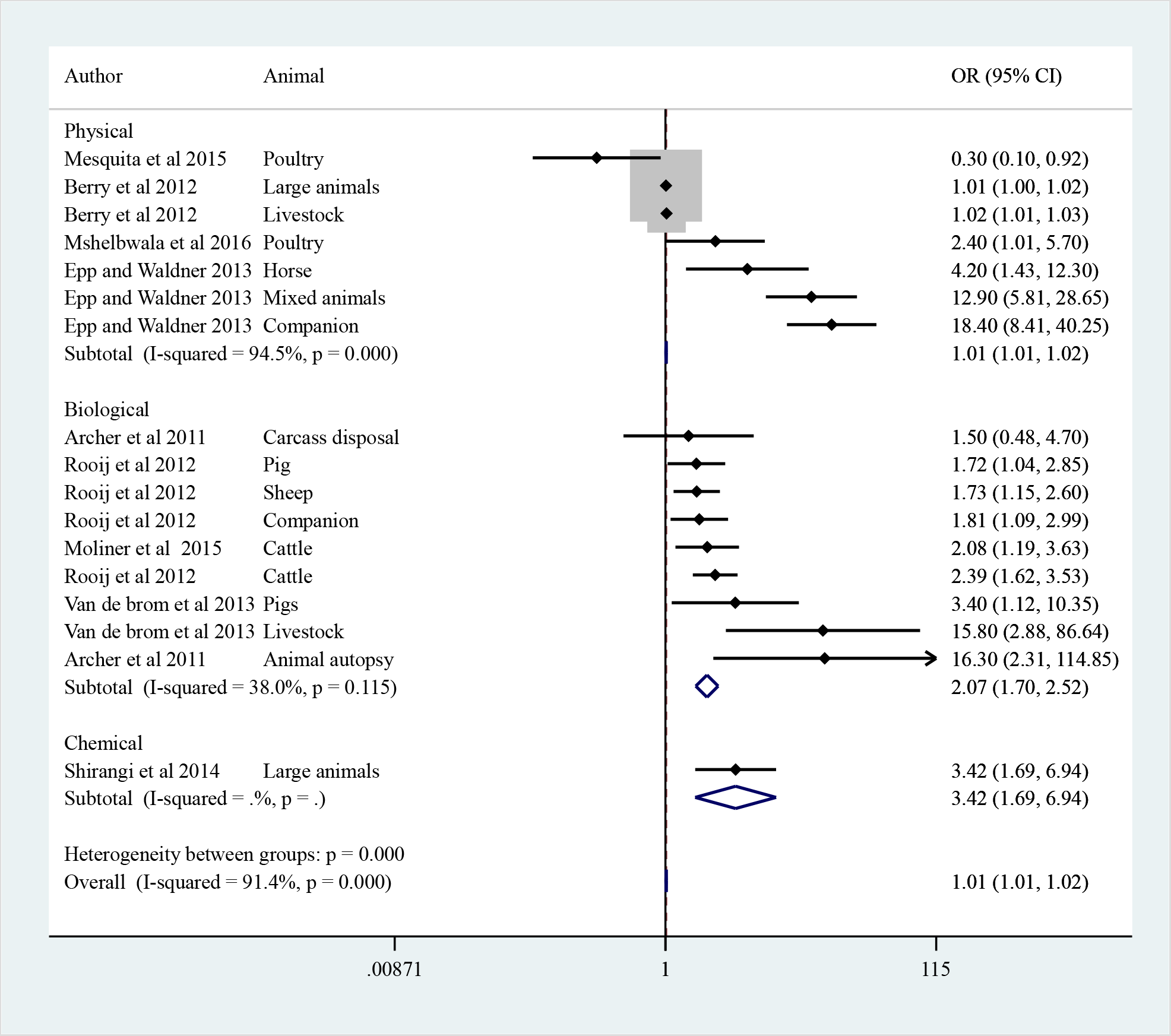
Forest plot of pooled odds ratio estimate of working or contact with animals as a risk factor for occupational hazards among veterinarians.

## 4. DISCUSSION

This work has shown that veterinarians and students are at high risk of diverse physical, chemical and biological hazards based on their work profiles – contact with animals. Data on occupational health and safety, particularly associated risks, are very limited in the developing countries, unlike in the developed ones. We were able to retrieve two papers from Nigeria and South Africa published between 2007 – 2017. This may be attributed to weak veterinary services and infrastructures in many developing countries, attitudinal issues to occupational health and safety, adherence to and implementation of existing policies, lack of disease/risk surveillance and monitoring systems at national or regional levels, poor laboratory capacitation for early detection and responses, poor disease/risk assessments, reporting and communication. These factors listed above are potential contributors to supposed underestimation of the hazards and may give the false impression that animal health-related workplace occupational hazards are not a problem in Africa. While this review focused on synthesizing and identifying literature on occupational hazards and associated factors in the veterinary setting, it has also revealed a gap with respect to the African continent.

Overall, veterinarians are exposed to diverse forms of hazards in their workplace. Significant physical hazards, particularly the needlestick injuries (NSI) (75.0%; 95% CI: 68.0–82.0%, p< 0.001) and cumulative traumatic disorder (CTD) (25%; 95% CI: 23.0–27.0%, p< 0.001) among veterinarians has been a source of worries for professionals due to their high prevalence [4,5,7,9,38]. NSI remains a major hazard and issue in the veterinary profession especially with regards to recapping of needles and consequent accidental needle prick [9]. Besides, frequent contact with and handling of large animals such as pigs, goats and sheep increased the risk of NSI in veterinary students[38]. The lack of in-depth understanding of animal behaviour (ethology), improper handling and disposal of needles, and poor or inadequate restraint techniques applied to these large animals predispose to increased risk of NSIs [38]. Emphasizing mitigation measures including the competent training and impartation of skills in the best practices relating to the handling, recapping and disposal of needles and sharps, as well as standard restraining techniques and facilities for animals must be the priority.

Cumulative traumatic disorders (CTDs) are chronic injuries marked with excessive pains in the musculoskeletal system due to overuse or repetitive strain from forceful motions or prolonged uncomfortable positions [39]. In veterinary practices, many activities are physically demanding and have increased potential for injury, especially when dealing with untamed or raging animals [11]. Veterinarians have ≈ 3 times higher likelihood of CTDs in their workplace when compared with the human doctors [40]. Similarly, large animal veterinarians were more predisposed to CTDs than counterparts in other practices and injuries more often occurred in the upper extremities and knees, cases which may become more severe with the pressure of time and work-related stress [11, 41].

Although few papers were obtained for chemical exposures and risk practices, the pooled estimate proportion was 7.0% (95% CI: 6.0–9.0%). It will appear that chemical hazards and quantitative exposure in the veterinary profession are less studied compared with physical and biological ones. Previous workers have suggested under-reporting, inconclusive data and the inability to recognize the causal link of hazard as possible reasons for the low prevalence of chemical risks observed in veterinary practices [41]. Veterinarians by practices undertake activities that expose them to chemicals and ionizing rays like radiations from radiography and radiology, pesticides and cytotoxic drugs during prophylaxis or therapy, anaesthetic gases during preparation for surgeries, and these can lead to life-threatening illnesses, disorders or complications as grave as cancer [42], reproductive health challenges like birth defects [8,12], disorders of the central nervous system, liver and kidney [42]. Our results indicated that female veterinarians more than male, especially those working in small animal practice are at higher odds of chemical exposures and attendant consequences [12]. Early notification of pregnancies and risk assessments at the workplace may be useful to mitigate the associated health risks.

Biological hazards, most particularly zoonoses remained the most emphasized, studied and documented risks based on our observation during this review. Significantly, these diseases still go under-reported in many countries and most especially in the developing ones where the risk of coinfections also exists. In this study, almost all of the reported studies originated from the developed economies (72.7% originating from Europe, North America and Australia alone). Approximately 61% of all human pathogens are zoonotic, and 75% of all emerging pathogens during the past decade have animal source. [43] The constant interaction and contact of veterinarians with animals categorize them as one of the high-risk groups for zoonoses and or emerging infectious diseases with the consequences of exposure ranging from the simple seroconversion to the disease with extremely variable symptomatic manifestations until the onset of irreversible sequelae or death [41]. This risk is especially problematic to people, such as companion animal owners and veterinary health workers who are immunocompromised [44]. in this work, zoonoses incidence ranged from 0.1 to 73.7% and some common zoonoses identified as threats with relatively high proportions were recorded for bartonellosis (31%), Q-fever (26.0%) and other viral infections (24%). Contact with or handling animals increases the risk of exposure to zoonoses by twice in veterinarians (OR = 2.07, 95% CI: 1.70–2.52, p < 0.001), emphasizing the importance of zoonoses in the veterinary profession. Mitigation strategies to reduce occupational-related zoonotic threats should be addressed through the implementation of policies and legal document that encourages compliance, improved mechanisms for effective risks assessment, communication and surveillance for domestic and wild animals and veterinary population. Targeted educational programme on zoonoses and EIDs and control strategies among veterinarians and students also becomes needed. We encourage more surveys on occupational health, documenting and reviewing the risks and the impact of zoonotic diseases on the veterinary profession would contribute to occupational risk prevention [45]. Development of risk assessment plans, health and safety guidelines, good practices, and mitigation systems to reduce workplace-related hazards must be carried out from training institutions to workplace postgraduation [38]. Finally, the fact that zero cases of MRSA and physical/psychological trauma were reported in students and the sections of students that experienced other forms of occupational exposures were in the clinical years were indications that practising veterinarians particularly based on years of experience on the job are at higher risks of occupational risks.

## CONCLUSIONS

We acknowledge this review may have experienced some limitations based on the number of search database (3) used and the omission of some important studies may have occurred.

However, this review has provided a better understanding of occupational health and safety status of veterinarians and gaps within the developing countries. We reported the veterinary field is challenged with diverse physical, chemical and biological hazards mainly zoonoses. Handling or being in contact with animals poses higher exposures to physical and biological hazards. Therefore, protecting the health and safety of veterinarians in their various workplaces becomes essential through collaborative efforts of all experts in the field, improved and efficient implementation of occupational health and safety policies, encouraging best preventive practices and safety standards, surveillance, targeted continuing educational programmes, risk assessment, reporting and communication. Furthermore, more studies may be required to quantify hazard exposures and impact on the health of veterinarians and students globally but more importantly in the developing countries where little studies have been conducted.

## Data Availability

The data that support the findings of this study are available from the corresponding author, upon reasonable request

## Authors contributions

OOA and MA generated the protocol with input from all authors. OOA, FOG, BB did the database search, abstract screening, and data extraction. BB did the data analysis. OOA drafted the manuscript with input from all the authors. FOF revised the manuscript critically for important intellectual content. All authors read and approved the final version of the manuscript to be published and agreed to be accountable for all aspects of the work.

## Acknowledgement

We thank the College Librarian, Dr K.A. Owolabi, College of Veterinary Medicine, Federal University of Agriculture Abeokuta, Ogun State, Nigeria for his technical support with the systematic reviews.

